# The acute effects of knee extension exercises with different contraction durations on the subsequent maximal knee extension torque for athletes with different strength levels

**DOI:** 10.1101/2022.04.12.22273776

**Authors:** Kaito Nakata, Takaaki Mishima

## Abstract

Individuals with high fatigue resistance against a high-intensity conditioning activity (CA) may be able to avoid experiencing significant fatigue and enhance their voluntary performance. We examined whether the optimal contraction duration of dynamic knee extension exercises to maximize subsequent voluntary performance varies depending on the strength level of an individual. The study participants were 22 male American college football players. Initially, all participants performed a 10-s maximal isometric knee extension exercise and were classified as stronger individuals (n = 8) and weaker individuals (n = 8) based on their relative muscle strength. Each group then performed three types of dynamic CA with different contraction durations (6 s [6-CA], 12 s [12-CA], and 18 s [18-CA]) in random order. To observe the time-course changes in post-activation potentiation and performance enhancement, the twitch torques induced by electrical stimulation and isokinetic knee extension torques at 180°/s were recorded before and after each CA. The twitch torque increased at 10 s (29.5% ± 9.3%) and 1 min (18.5% ± 6.8%) after 6-CA for the stronger individuals (*p* < 0.05). However, no post-activation potentiation was induced in the weaker individuals in either protocol. Voluntary performance increased at 4 (7.0% ± 4.5%) and 7 (8.2% ± 4.3%) min after 18-CA for stronger individuals (*p* < 0.05). However, there was no post-activation performance enhancement in either protocol for weaker individuals. Thus, CA with a relatively long contraction duration was optimal to maximize the subsequent voluntary performance for stronger individuals. It remains unknown whether CAs performed with relatively short or long contraction durations were optimal for weaker individuals.

## Introduction

The twitch torque evoked by a single electrical stimulus is increased transiently after a maximal or submaximal conditioning activity (CA) [1,2]. This phenomenon is referred to as post-activation potentiation (PAP). The primary mechanism of PAP is the phosphorylation of myosin regulatory light chains (P-MRLC) during a CA [3,4]. P-MRLC transiently increases the sensitivity of actin–myosin interactions to a given amount of Ca^2+^. As a result, the force-generating capability of the muscle is enhanced [5,6]. Several previous studies have reported that a CA can improve not only the twitch torque evoked by electrical stimulation but also the subsequent voluntary performance [7–11] called the post-activation performance enhancement (PAPE) [12]. Tillin and Bishop [6] described P-MRLC as one of the major mechanisms of PAPE.

The potentiating effect and fatigue associated with a CA coexist [13]. Therefore, an increase in the voluntary performance after a CA appears to depend on the net balance between fatigue and potentiation [6]. Specifically, voluntary performance may decrease if the effects of fatigue are dominant but increase if those of potentiation dominate or may remain unchanged if the effects of fatigue and potentiation are at similar levels. Therefore, in situations where coaches are attempting to employ a CA to sufficiently enhance the subsequent voluntary performance, they need to consider both inducing the potentiating effect and suppressing the fatigue associated with a CA as much as possible.

The fact that the total contraction duration of a CA plays a crucial role in maximizing PAPE deserves more attention. If the total contraction duration of a CA is too short, the mechanisms associated with the potentiating effect may not be triggered. Seitz et al. [14] examined the effect of the total contraction duration of isokinetic dynamic knee extension exercises on PAP and PAPE. They observed that a minimum total contraction duration must be reached during a CA to trigger the mechanisms responsible for PAP and PAPE. However, if the total contraction duration of a CA is too long, the potentiating effect induced by the CA may be counteracted by significant fatigue. Hamada et al. [15] used a fatiguing protocol of 16 sets of 5-s maximal voluntary contraction (MVC) and induced twitch torque between each set to observe the time-course changes in PAP. Their results indicated that the fatigue accumulated by the continuous MVCs counteracted the potentiating effect of P-MRLC. Therefore, a CA with an excessively long total contraction duration that induces significant fatigue would not be suitable for enhancing the subsequent voluntary performance. It follows that to effectively induce PAPE, it is necessary to identify the optimal total contraction duration of a CA that can sufficiently trigger the potentiating effect while minimizing the fatigue as much as possible.

Interestingly, previous studies have suggested that the magnitude of PAP [1] and fatigue [16–18] after a high-intensity CA vary depending on the strength level of an individual. Typically, stronger individuals (SI) have higher fast-twitch fiber content than weaker individuals (WI) [19,20]. It should be noted that the activity of myosin light chain kinase, the enzyme that triggers P-MRLC, is more prevalent in fast-twitch fibers than in slow-twitch fibers, resulting in P-MRLC occurring more prominently in fast-twitch fibers [3]. Therefore, during a CA, SI with a higher content of fast-twitch fiber may cause greater P-MRLC than that caused by WI, resulting in higher PAP [6]. In addition, it must be noted that the fatigue resistance against a high-intensity CA is greater in individuals with higher relative muscle strength [16–18]. Previously, Miyamoto et al. [17] confirmed that after a 5-s maximal isometric knee extension exercise as a CA, the dynamic maximal knee extension torque increased between 1 and 3 mins but did not increase immediately after the CA. Nevertheless, after 12 weeks of high-intensity knee extension training, the dynamic maximal knee extension torque significantly increased, even immediately after the CA. The main reason for the earlier realization of PAPE after the intervention is that the individuals developed resistance to fatigue by chronic high-intensity resistance training, suppressing the remarkable fatigue associated with a high-intensity CA. This result implies that SI have a greater fatigue resistance to a high-intensity CA compared with that of WI. Given that the extent of PAP and fatigue after a high-intensity CA vary depending on the strength level of an individual, we hypothesized that the optimal contraction duration of a CA to maximize PAPE may differ between SI and WI.

If a high-intensity CA with a relatively long contraction duration (i.e., approximately 20 s) is performed, it is likely that PAPE is not achieved because significant fatigue may counteract the potentiating effect. However, SI with high fatigue resistance against a high-intensity CA may have the potential to suppress the development of significant fatigue and enhance the subsequent voluntary performance, even in the CA with a relatively long total contraction duration. Furthermore, in SI with a higher number of fast-twitch fibers, the PAP mechanism may be triggered to a great extent by the performance of CA with a long total contraction duration, resulting in a greater extent of PAPE. On the other hand, when a high-intensity CA with a relatively short contraction duration (i.e., approximately 5 s) is performed, the magnitude of the potentiating effect is expected to be lower than that observed when a high-volume CA is performed. However, a short contraction duration may produce less fatigue and allow the potentiating effect to dominate, even in WI with relatively low fatigue resistance. Thus, the optimal contraction duration of a CA to maximize PAPE may vary depending on the strength level of an individual. A recent meta-analysis reported that SI tend to show more PAPE when a high-intensity CA is performed at maximal repetition than at sub-maximal repetition, whereas the opposite may be observed for WI [21]. However, to the best of our knowledge, no studies have examined in detail whether the optimal contraction duration of a high-intensity CA differs among individuals with different muscle strength levels.

Therefore, this study aimed to verify whether the optimal contraction duration of a high-intensity CA to maximize PAPE varies depending on the strength level of an individual. We hypothesized that the CA with a relatively long contraction duration (i.e., approximately 20 s) would be optimal for SI, while the CA with a relatively short contraction duration (i.e., approximately 5 s) would be optimal for WI.

## Materials and methods

### Participants

Twenty-two male American college football players participated in this study. All participants had participated in regular resistance training for at least 6 months. During the experimental period, they were instructed to continue their daily routines and to not change their physical activity level and food and fluid intakes. Moreover, they were asked to avoid consuming depressive (e.g., alcohol) or ergogenic (e.g., coffee) substances 24 h prior to the experimental sessions. They had been free from musculoskeletal injuries for at least 1 year before the study. They were provided oral and written explanations of the purpose, contents, and procedures of this study prior to the experiment, and the experiment was conducted after obtaining their consent. This study was approved by the Ethics Review Committee of Osaka University of Health and Sport Sciences (approval number: 21-2). The study was conducted in conformity with the policy statement regarding the use of human participants by the Declaration of Helsinki.

### Study design

This study was conducted over four sessions for each participant. In the first session, the participants were grouped into SI or WI based on their relative muscle strength, which was calculated by dividing the isometric maximal knee extension torque by the body weight. For the latter three sessions (i.e., experimental sessions), the particiants who were classified as SI or WI in the first session were asked to participate. Each experimental session was conducted at least 24 h apart. All sessions were conducted between 10:00 AM and 3:00 PM in an indoor laboratory. To determine the optimal contraction duration of a CA to maximize PAPE, each participant performed three types of dynamic CAs with different total contraction durations (6 s [6-CA)], 12 s [12-CA], and 18 s [18-CA]) in random order. The joint angular velocity of each CA was set at 60°/s. To observe the time-course changes in PAP and PAPE, before and after each CA, the twitch torques induced by electrical stimulation and voluntary knee extension torques at 180°/s were recorded. The repeatability (intraclass correlation coefficient [ICC]) of the peak torque of maximal voluntary isokinetic knee extension and twitch contraction was investigated in our preliminary study. The ICC values were 0.962 (0.897–0.989) and 0.998 (0.995–1.000) for voluntary knee extension peak torque and twitch peak torque, respectively. In all sessions, the participant was required to rest for 10 min after arriving at the laboratory to eliminate the effect of PAP induced by traveling to the laboratory. This rest period is sufficient to dissipate the effects of PAP [1,2]. The participants sat on a dynamometer (Biodex System 4; Sakai Medical Instrument, Tokyo, Japan) and were tightly secured to the seat using two crossover seatbelts and a waist harness. The lever arm of the dynamometer was attached 2–3 cm above the lateral malleolus with a strap. The rotation axis of the knee was aligned with the axis of the motor. The peak torque of voluntary knee extension and twitch contraction of the dynamometer was analog-to-digital converted (Power Lab/8SP; ADInstruments, Bella Vista, NSW, Australia) and stored on a personal computer software (LabChart 6 Japanese; ADInstruments). Data were filtered with a cut-off frequency of 12 Hz. In addition, MacIntosh et al. [22] indicated a possibility that the learning and familiarization effect of the performance task could affect PAPE. Therefore, in the present study, we conducted a task-specific warm-up session, based on the study by Seitz et al. [14], before each experimental session to eliminate the possibility of the learning and familiarization effect.

### Procedure

#### Classification of SI and WI

To classify SI and WI, all participants performed a 10-s maximal voluntary isometric knee extension exercise and were consistently encouraged to contract “as forcefully and as fast as possible.” To observe the relationship between the muscle strength levels and the magnitude of PAP, a twitch torque was induced by electrical stimulation before and immediately after a 10-s isometric knee extension exercise. The extent of PAP induced by isometric knee extension exercise (%PAP_MVC_) was calculated as follows:

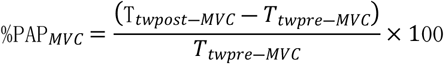

where *T*_*twpre―MVC*_ and T_*twpost―MVC*_ are the maximal twitch torques obtained before and immediately after the isometric knee extension exercise, respectively. From an initial sample of 22 men, 8 participants demonstrating the highest and 8 demonstrating the lowest relative muscle strength were selected and classified as SI and WI, respectively. The other 6 participants were not classified into any of the groups and did not participate in any further experiments.

#### Electrical stimulation procedure

To evoke quadriceps muscle twitches, an electrode pad consisting of a 10 × 20 cm aluminum foil with an adhesive conductor (SR-4080; Minato Medical Science, Osaka, Japan) attached and covered with kitchen paper was prepared. Two electrode pads were wetted with water and attached to the upper and lower anterior thigh to cover the entire quadriceps muscle of the dominant leg. The participant was then placed on the dynamometer seat with the knee and hip joints fixed at 90° and 80°, respectively. The electrode pads were connected to a high-voltage constant-current stimulator (Model DS7AH; Digitimer Ltd, Hertfordshire, UK) via an output cable (D185-HB4; Digitimer Ltd, Hertfordshire, UK). The stimulus intensity was set at 120% of the maximal intensity [15,23].

#### Task-specific warm-up sessions

An overview of the task-specific warm-up session is shown in Fig 1A. After we determined the stimulation intensity, the participants performed two isokinetic knee extension exercises at 180°/s at 20%, 40%, 60%, and 80% of their perceived maximal torque at 45-s intervals. Isokinetic knee extensions at 100% of the maximal torque were then performed “as fast and as hard as possible” every minute until the peak torque production in three consecutive contractions differed by less than 2%. According to a previous study [14], the angular velocity of the performance test was set at 180°/s. In addition, the range of motion of the knee was between 110° and 20° (0°, full extension) in concentric contractions. The participant performed a maximal isokinetic knee extension exercise over the entire range of motion until the dynamometer mechanically stopped.

**Fig 1.**
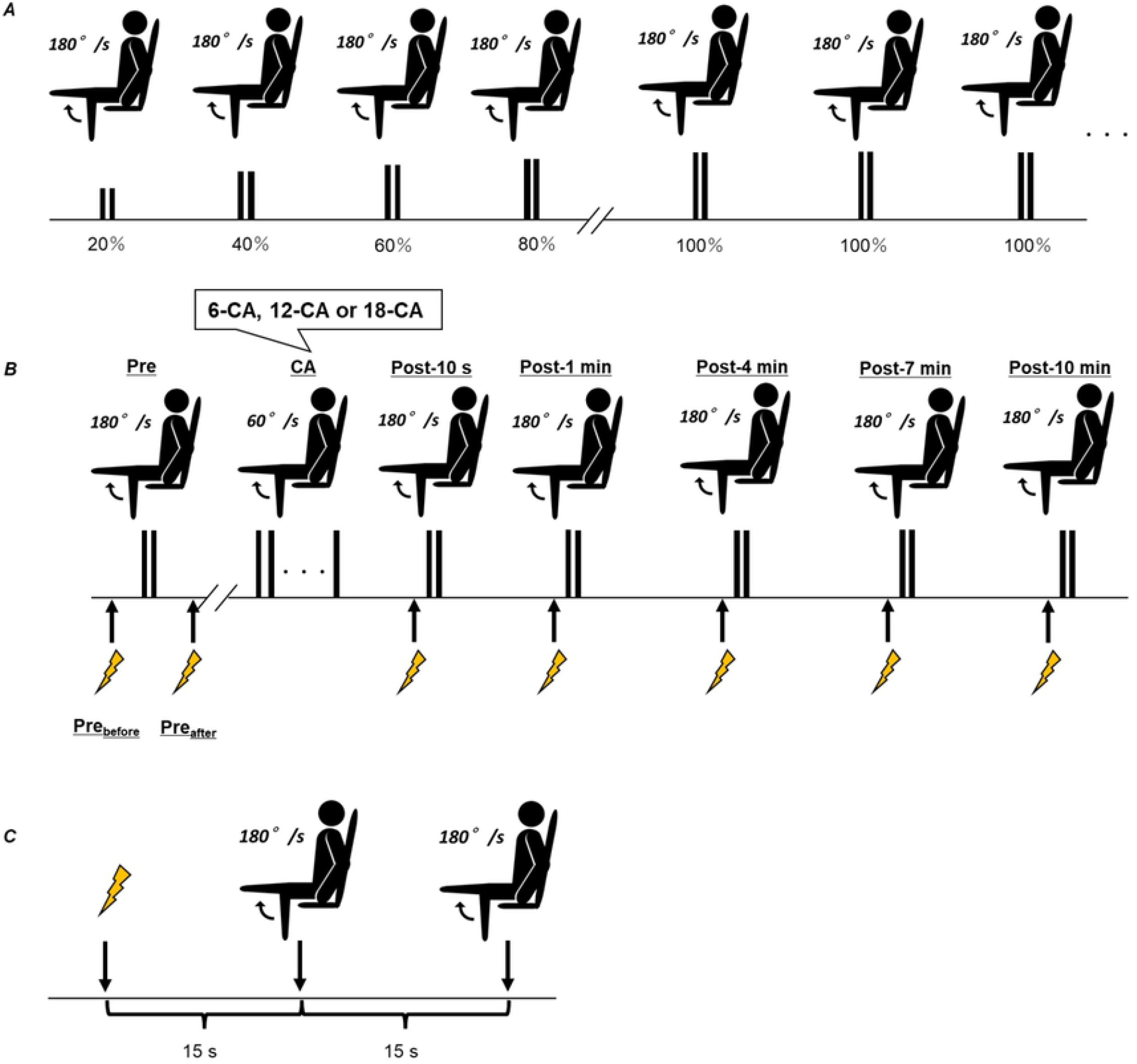
Experimental overview. (*A*) Task-specific warm-up procedure (*B*) Experimental procedure for the 6-CA, 12-CA, and 18-CA protocols. (*C*) The details of the test protocol after each CA. CA, conditioning activity.

#### Experimental session

After the task-specific warm-up session, a 10-min rest period was provided before the experimental session. An overview of the experimental session and the posttest protocol are shown in Fig 1B and 1C, respectively. In the experimental session, the responses to one isometric twitch torque (Pre_before_) and two isokinetic knee extension exercises at 180°/s followed 5-s later by one isometric twitch (Pre_after_) were recorded before each participant performed each CA. The last stimulation was used to examine the possible potentiating effect of a set of two maximal isokinetic knee extensions at 180°/s. Following a rest period of approximately 90-s after Pre_after_, one of the three dynamic CAs with different contraction durations (6-CA, 12-CA, or 18-CA) was performed. The posttest was completed 10 s (Post-10 s) and 1 (Post-1 min), 4 (Post-4 min), 7 (Post-7 min), and 10 (Post-10 min) min after each CA (Fig 1B). The posttest protocol consisted of a single stimulation and two isokinetic knee extension exercises at 180°/s (Fig 1C). The range of motion of the knee was the same as in the task-specific warm-up session. The knee extension resulting in the highest voluntary peak torque at each time point (i.e., Pre, Post-10 s, Post-1 min, Post-4 min, Post-7 min, and Post-10 min) was selected for further analysis. After the first knee extension exercise, the lever arm of the dynamometer was manually returned to the starting position (i.e., 110° of knee flexion) and then the second knee extension exercise was performed. The extent of PAPE (%PAPE) was calculated as follows:

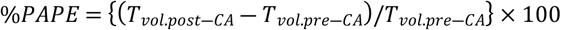

where T_*vol.pre*-*CA*_ and *T*_*vol.post*-*CA*_ are the maximal knee extension torques during isokinetic knee extension exercises before and after each CA, respectively. The extent of PAP measured in the experimental session was expressed as %PAP. Among them, the %PAP observed in two maximal knee extension exercises during the pretest was calculated as follows:

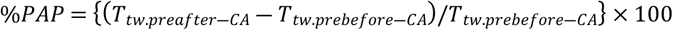

where *T*_*tw.prebefore―CA*_ and *T*_*tw.preafter―CA*_ are the maximal twitch torques induced before and after the two isokinetic knee extension exercises during the pretest, respectively. Finally, the %PAP observed after each CA was calculated as follows:

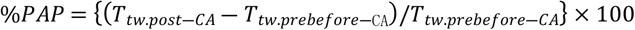

where *T*_*tw.post―CA*_ is the maximal twitch torque induced at each time point after each CA.

### CA protocols

Based on a previous study by Seitz et al. [14], the dynamic CAs employed in this study were isokinetic knee extension exercises at 60°/s. The numbers of contractions for 6-CA, 12-CA, and 18-CA were 4, 8, and 12, respectively. The total contraction duration was calculated as follows:

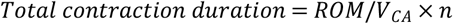

where *ROM* is the range of motion of the knee extensions performed during each CA (i.e., 90°), *V*_*CA*_ is the angular velocity of the knee extensions (i.e., 60°/s), and *n* is the number of knee extensions performed during each CA.

### Statistical analysis

Descriptive data are presented as mean ± standard deviation. A two-tailed, unpaired *t*-test was used for comparisons between the SI and WI groups. Pearson’s correlation coefficients (*r*) were calculated and classified as small (0.1 *≤ r* < 0.3), moderate (0.3 *≤ r* < 0.5), and large (*r ≥* 0.5). One-way analysis of variance (ANOVA) with repeated measures was performed to compare the voluntary knee extension torques produced during the last three knee extensions of the task-specific warm-up session and during the pretest knee extension to determine whether the task-specific warm-up was completed. For %PAP, a two-way ANOVA (time [Pre_before_, Pre_after_, Post-10 s, Post-1 min, Post-4 min, Post-7 min, Post-10 min] × protocol [6-CA, 12-CA, 18-CA]) with repeated measures was used. Similarly, for %PAPE, two-way ANOVA (time [Pre, Post-10 s, Post-1 min, Post-4 min, Post-7 min, Post-10 min] × protocol [6-CA, 12-CA, 18-CA]) with repeated measures was used. When a significant interaction was found, the Bonferroni post-hoc test was used to examine the time-course changes in each CA protocol. Effect sizes were estimated by calculating the partial eta-squared (*η*^*2*^) values (small: 0.01–0.059, moderate: 0.06–0.137, and large: > 0.138). For pairwise comparisons, effect size was determined by Cohen’s *d* (small: > 0.2, moderate: > 0.5, large: > 0.8). Statistical analyses were performed using statistical software (SPSS Statistics 27; IBM Japan, Tokyo, Japan). The significance level for all comparisons was set at *p* < 0.05.

## Results

### Comparison of SI and WI and the relationship between % PAP_MVC_ and muscle strength

An unpaired *t*-test showed a significant difference between SI and WI in absolute (*p* < 0.001 *d* = 2.427) and relative (*p* < 0.001, d = 3.434) muscle strength. There was also a significant difference between SI and WI in %PAP_MVC_ (*p* = 0.004, d = 1.806) (Table 1). A significant relationship was found between %PAP_MVC_ and the absolute (*r* = 0.550, *p* = 0.008) and relative (*r* = 0.570, *p* = 0.006) muscle strength for all the participants (Fig 2).

**Table 1.** Physical characteristics, MVC, twitch torque, and %PAPMVC in the whole group and SI and WI subgroups.

|  | Whole (n = 22) | SI (n = 8) | WI (n = 8) |
| --- | --- | --- | --- |
| Age (year) | 19.7 ± 1.2 | 19.9 ± 1.6 | 19.8 ± 1.0 |
| Training Age (year) | 3.5 ± 1.2 | 3.5 ± 1.1 | 3.5 ± 1.7 |
| Height (m) | 171.5 ± 6.1 | 172.5 ± 6.5 | 169.1 ± 5.7 |
| Weight (kg) | 74.6 ± 10.6 | 72.3 ± 10.4 | 74.8 ± 13.2 |
| MVC (N·m) | 274.7 ± 68.2 | 337.6 ± 41.6* | 204.3 ± 37.3 |
| MVC/Weight (N·m·kg<sup>-1</sup>) | 3.7 ± 0.9 | 4.7 ± 0.5* | 2.4 ± 1.0 |
| pre-MVC (N·m) | 44.6 ± 8.9 | 43.7 ± 6.0 | 47.3 ± 9.4 |
| post-MVC (N·m) | 71.3 ± 10.6 | 75.3 ± 8.1 | 67.1 ± 12.9 |
| %PAP<sub>MVC</sub> (%) | 63.2 ± 29.1 | 73.5 ± 11.2* | 43.4 ± 22.4 |

Values are presented as means ± standard deviation. MVC, maximal voluntary contraction; pre-
MVC, twitch torque before MVC; post-MVC, twitch torque after MVC; SI, stronger individuals;
WI, weaker individuals. The statistical difference is set at $p < 0.05$ : difference versus WI\*

**Fig 2.**
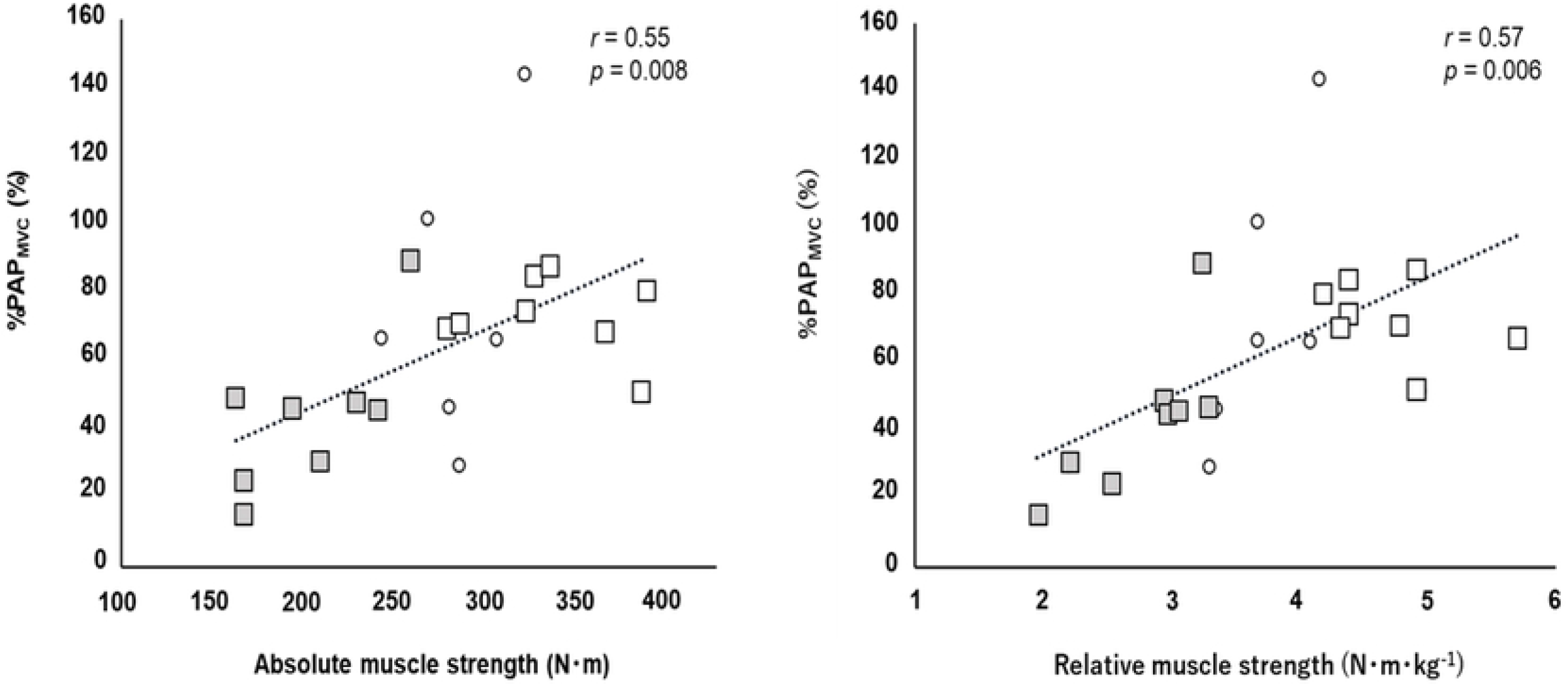
Correlation between %PAP_MVC_ and absolute (*A*) and relative (*B*) muscle strength. Filled squares, open squares, and open circles represent the weaker, stronger, and intermediate levels of the relative muscle strength, respectively

### Task-specific warm-up session

In the task-specific warm-up session for each protocol, the numbers of knee extensions for 6-CA, 12-CA, and 18-CA were 4.4 ± 1.7, 4.9 ± 2.7, and 8.6 ± 4.8 for SI, and 7.0 ± 4.6, 7.3 ± 4.8, and 7.8 ± 1.4 for WI, respectively. Fig 3 shows no significant difference between the maximal voluntary knee extension torque during the last three knee extensions in the task-specific warm-up session and the maximal voluntary knee extension torque during the pretest in each CA protocol. The fact that no significant difference was observed between the knee extension torques in the condition in which sufficient practice was conducted and in the pretest indicates that the task-specific warm-up session was sufficiently completed before conducting each CA and the effects of learning and familiarization on PAPE were minimized.

**Fig 3.**
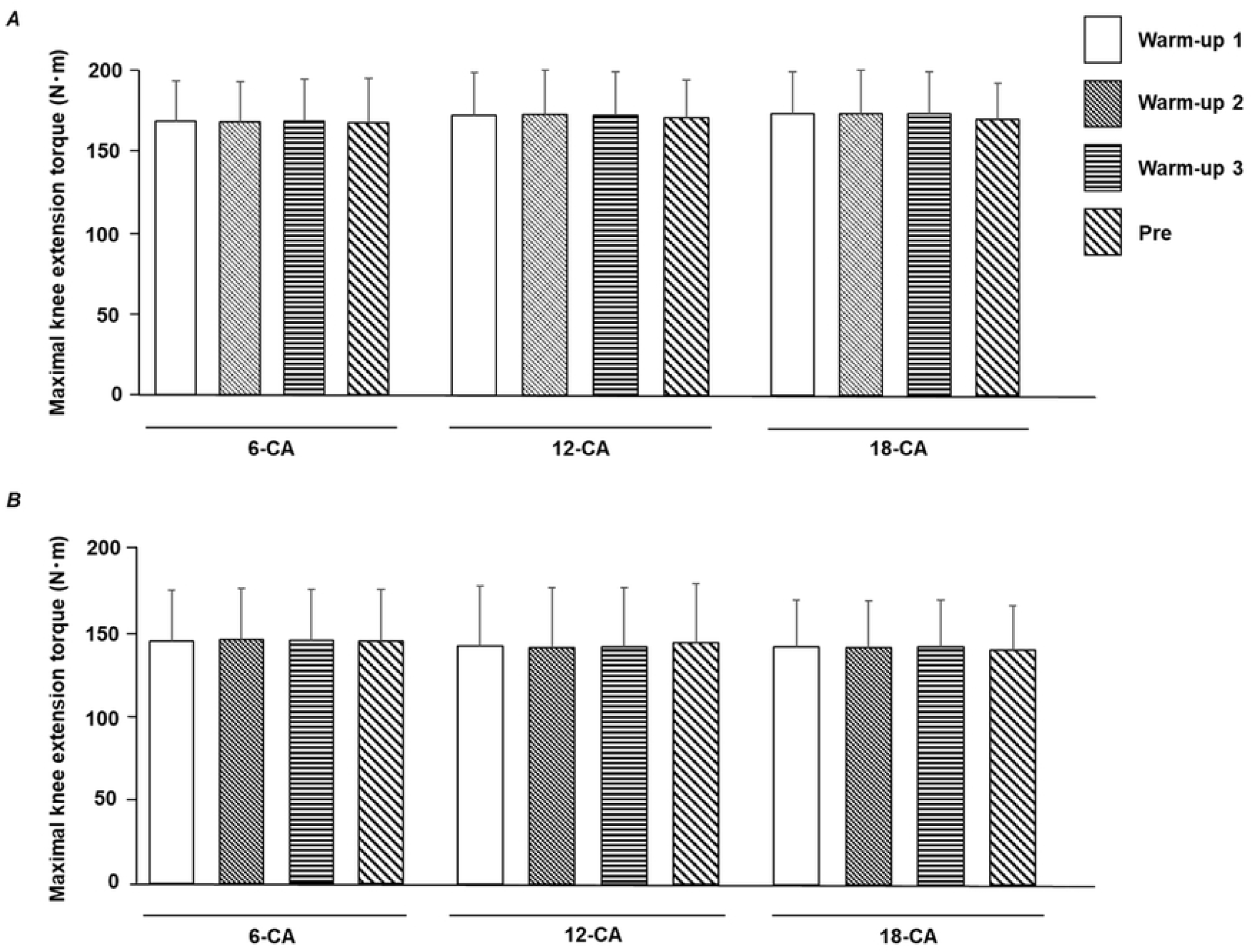
The task-specific warm-up session was adequately performed. Maximal voluntary knee extension torques produced during the last three knee extensions at the task-specific warm-up session (warm-up 1, warm-up 2, and warm-up 3) and the knee extension during the pretest for (*A*) stronger individuals (SI) and (*B*) weaker individuals (WI). CA, conditioning activity.

### Twitch torque

For the twitch torque, the two-way ANOVA revealed that the time × protocol interaction was significant for SI (*p* < 0.001, *η*^*2*^ = 0.404). Further analyses showed that in all protocols, the peak twitch torque after a set of two isokinetic knee extensions (Pre_after_) was significantly potentiated (6-CA, 15.7% ± 6.3%, *p* = 0.004, d = 1.068, 95% confidence interval (CI): 10.4–21.0; 12-CA, 14.1% ± 8.4%, *p* = 0.045, d = 0.417, 95% CI: 7.0–21.2; 18-CA, 11.7% ± 3.6%, *p* = 0.001, d = 1.263, 95% CI: 8.7–14.7), indicating that two isokinetic knee extensions induce PAP. Moreover, for only 6-CA, the values of Post-10 s (29.5% ± 9.3%, *p* = 0.001, d = 1.714, 95% CI: 21.7–37.3) and Post-1 min (18.5% ± 6.8%, *p* = 0.003, d = 1.101, 95% CI: 14.8–24.2) were significantly higher than the Pre_before_ value (Fig 4A). Fig 4B shows the significant interaction effect for the twitch torques for WI (*p* < 0.001, *η*^*2*^ = 0.369). However, further analyses revealed no significant difference between the baseline value (Pre_before_) and the values at the other time periods for each CA protocol.

**Fig 4.**
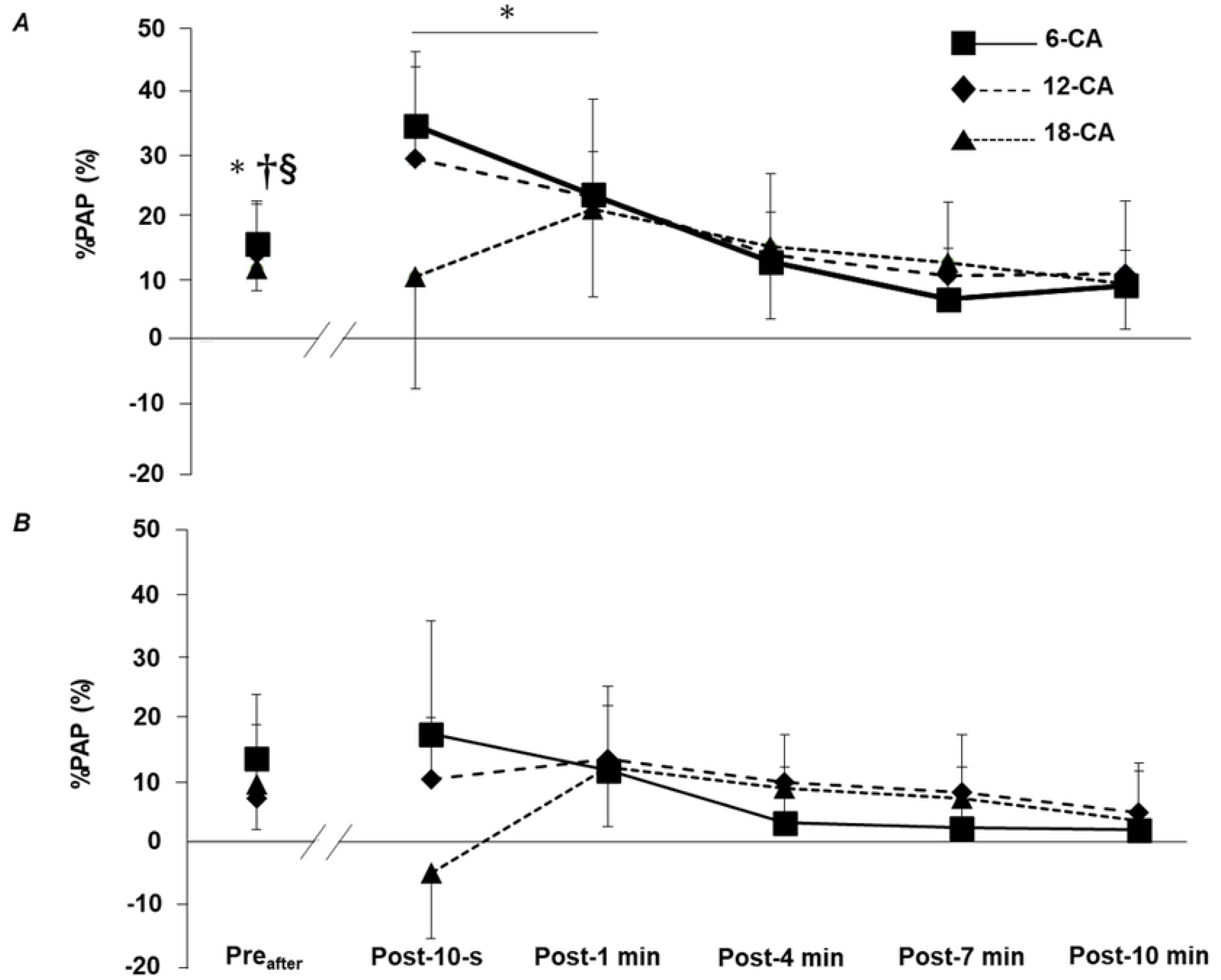
Time-course changes in %PAP after each CA for (*A*) stronger individuals (SI) and (*B*) weaker individuals (WI) CA, conditioning activity. Values are presented as means ± standard deviation. The statistical difference is set at *p* < 0.05: difference from Pre_before_ in 6-CA*, 12-CA^†^, and 18-CA^§^.

### Voluntary knee extension torque

Fig 5A shows the significant interaction effect for %PAPE for SI (*p* < 0.001, *η*^*2*^ = 0.377). For the 18-CA protocol, further analyses revealed that the values of Post-4 min (7.0% ± 4.5%, *p* = 0.047, d = 0.494, 95% CI: 3.2–10.7) and Post-7 min (8.2% ± 4.3%, *p* = 0.016, d = 0.568, 95% CI: 4.6– 11.8) were significantly higher than the baseline value (Pre). In contrast, there was no significant interaction for %PAPE for WI (*p* = 0.057, *η*^*2*^ = 0.215). In addition, the results of %PAPE for each participant after each CA protocol are shown in Fig 6.

**Fig 5.**
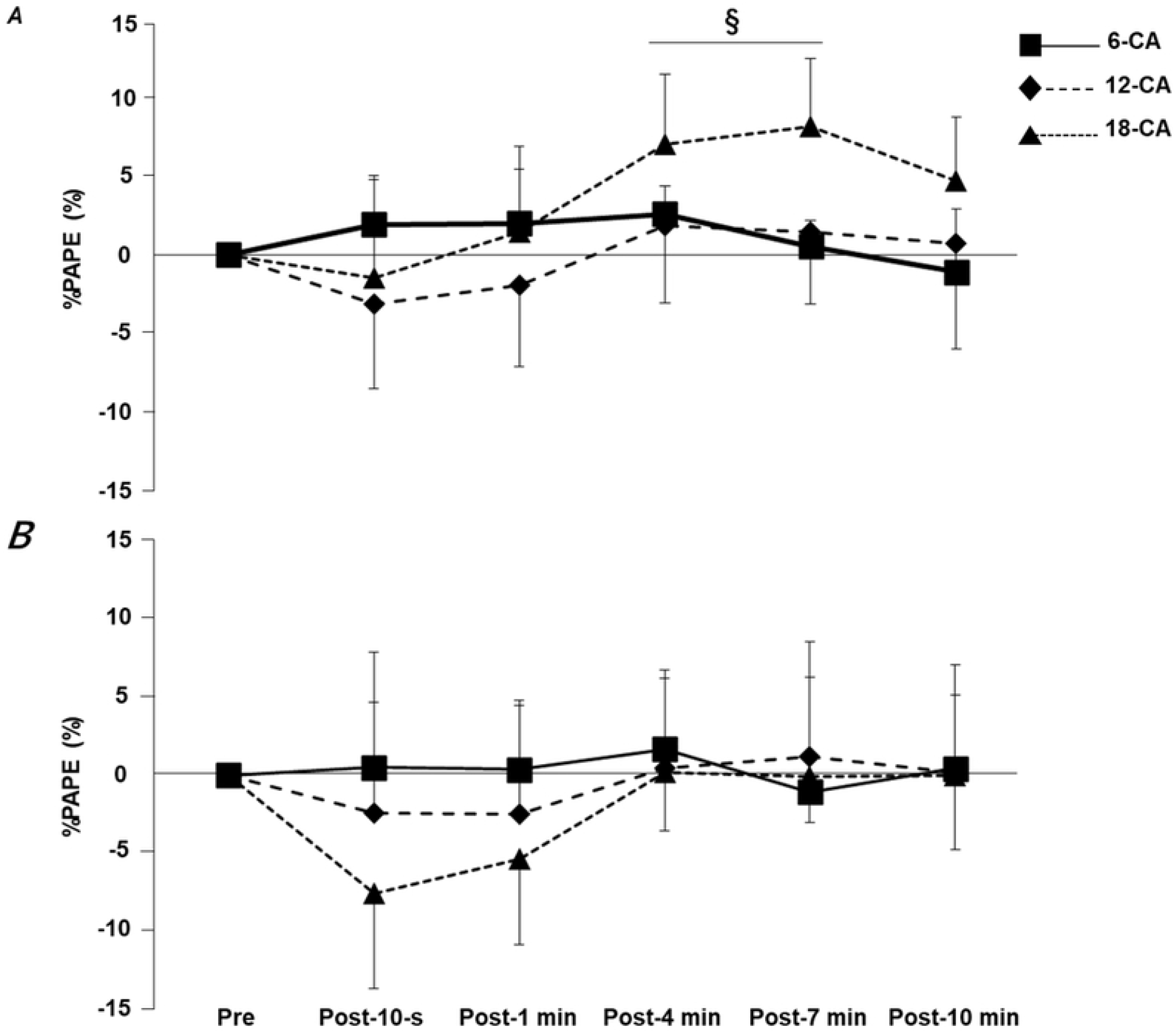
Time-course changes in %PAPE after each CA for (*A*) stronger individuals (SI) and (*B*) weaker individuals (WI) CA, conditioning activity. Values are means ± standard deviation. The statistical difference is set at *p* < 0.05: difference from Pre in 18-CA^§^

**Fig 6.**
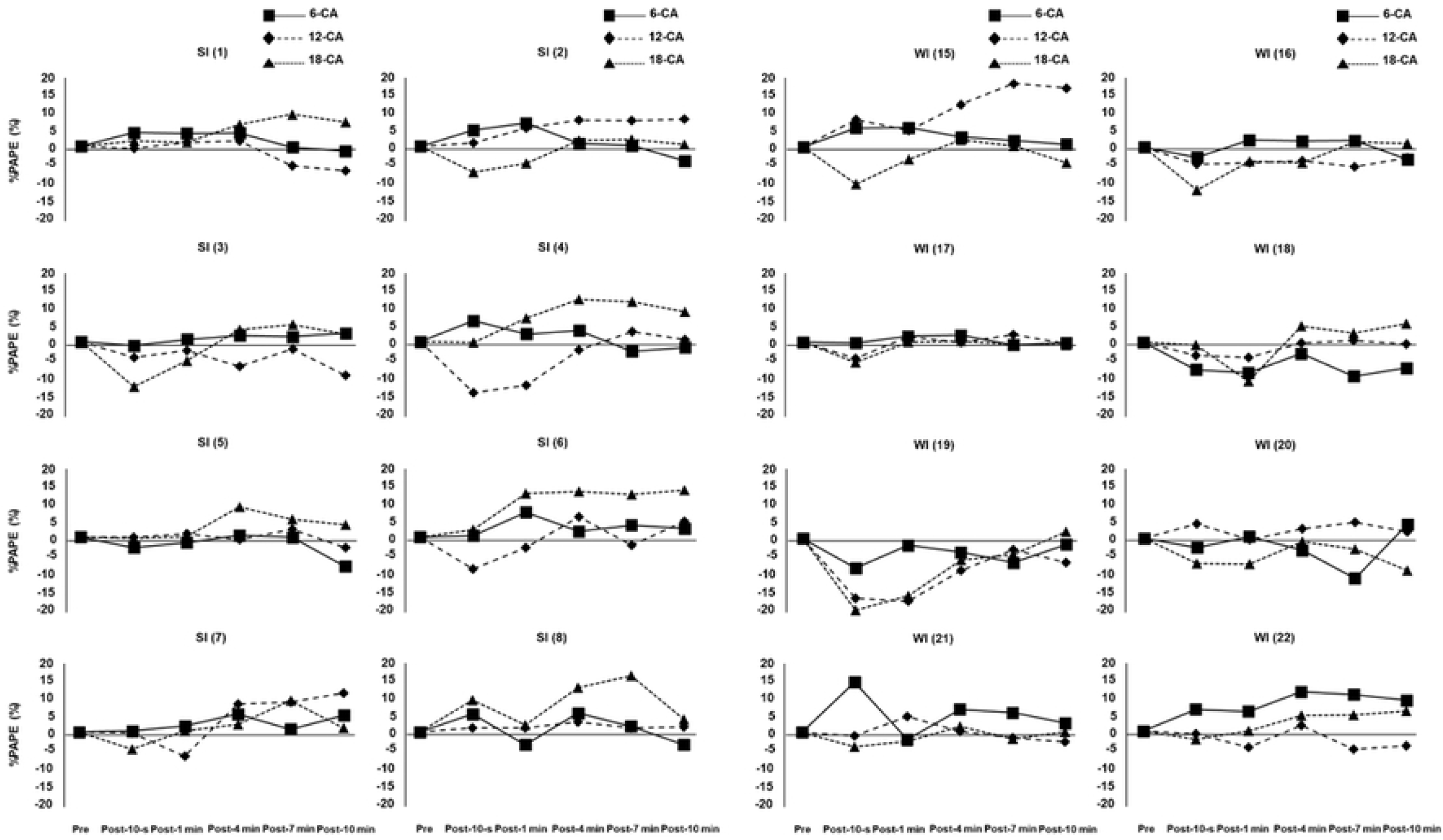
Time-course changes in %PAPE after each CA for each individual. The number in parentheses indicates how high the individual’s relative muscle strength was from the top among all participants. SI, stronger individuals; WI, weaker individuals; CA, conditioning activity

## Discussion

This study aimed to determine whether the optimal contraction duration of a CA to maximize PAPE differs among athletes with different muscle strength levels. The results obtained in the present study suggest that the strategies for maximizing PAPE may differ depending on the strength level of an individual.

In the present study, we observed that SI improved the voluntary performance only in the 18-CA protocol (Fig 5A), and when the results of PAPE were examined for each individual, most participants (i.e., individuals numbered 1, 3, 4, 5, 6, and 8) enhanced their voluntary performance after the 18-CA protocol more than after the other two protocols (Fig 6). These facts suggest that the CA with a relatively long contraction duration was optimal for SI. However, it should be noted that PAP had already disappeared at the time when PAPE was observed in the 18-CA protocol (i.e., Post-4 min, Post-7 min; Fig 4A), which indicates that PAP was not involved in the mechanism of PAPE in the 18-CA protocol. Previous studies have also confirmed the presence of PAPE when PAP has already disappeared [24,25]. Prieske et al. [24] reported that the jump performance was significantly improved after the CA, but PAP was not observed at that time and concluded that PAP was not directly linked to the voluntary performance enhancements after the CA. Moreover, Thomas et al. [25] observed a significant increase in the countermovement jump height 8 min after performing the squat exercise, but PAP was no longer present at that time. Blazevich et al. [26] indicated that factors other than PAP, such as an increase in the muscle temperature, a change in the central drive capacity, and the learning and familiarization effect, may contribute to acute voluntary performance enhancements. De Ruiter and De Haan [27] reported that parameters such as the maximal power production improved as muscle temperature increased. Therefore, it is possible that increasing the muscle temperature associated with a CA would temporally enhance the subsequent voluntary performance. Furthermore, González-Alonso et al. [28] showed that the quadriceps muscle temperature continued to increase during a 3 min voluntary knee extension exercise. Based on this study, it is likely that the 18-CA protocol, which had the longest total contraction duration, increased the muscle temperature more than that observed with the other two protocols. Therefore, the fact that PAPE was observed only in the 18-CA protocol for SI may have been related to the difference in the degree of increase in the muscle temperature. In addition, Folland et al. [29] confirmed that the H-wave amplitude, normalized by the maximal amplitude of the M-wave, increased 5–11 min after the isometric knee extension exercise. This result indicates that a CA has the potential to increase the excitability of the motoneuron pool. Therefore, in the present study, it is possible that the CA increased the excitability at the spinal level, resulting in the increased recruitment of higher-order motor units. Furthermore, MacIntosh et al. [22] referred to the possibility that the learning and familiarization effects regarding the task performance could affect the acute enhancement of the voluntary performance. However, in the present study, the participnts completed the task-specific warm-up session to the point where no further improvement in the voluntary knee extension torque production could be achieved. Thus, it is unlikely that the learning and familiarization effects on task performance influenced the PAPE in this study. Collectively, factors other than PAP and the learning and familiarization effects, such as the increase in the muscle temperature and change in the central drive capacity, may be involved in the improvement of the voluntary knee extension performance observed in the 18-CA protocol for SI. However, since these factors were not investigated in our study, we cannot conclude whether these factors were involved in the potentiation of the knee extension performance in the 18-CA protocol for SI.

We should note why SI was unable to improve the voluntary performance in the condition in which PAP was present in the 6-CA protocol (i.e., Post-10 s, Post-1 min, Fig 4A and Fig 5A). The finding regarding the absence of the potentiation of the voluntary performance despite the occurrence of the maximal twitch potentiation has been confirmed in other studies. Fukutani et al. [23] researched the effect of a 6-s isometric ankle plantar flexion exercise on PAP and PAPE and found that isometric twitch torque and isokinetic voluntary plantar flexion torque at 180°/s significantly increased by 62.7% and 6.4%, respectively, immediately after the CA. However, 1 min after the CA, although PAP remained significantly higher than that at baseline, its potentiation effect decreased to 30.3%, and the voluntary performance was no longer significantly different from that at baseline. Importantly, sensitivity to the potentiation effect of a CA differs between the twitch torque and maximal voluntary torque [4]. The Ca^2+^ concentration is much higher in the maximal voluntary concentric contraction (multiple impulses) than in the twitch contraction (single impulse). In addition, the extent of increase in the torque by a CA becomes smaller when the Ca^2+^ concentration is higher because, in this case, several cross-bridge attachments already exist [30]. In this context, Zimmermann et al. [31] noted that PAP must be extremely high to contribute to an increase in voluntary performance. Therefore, it is reasonable to conclude that the potentiation effect observed after 6-CA was insufficient to enhance the voluntary performance to the point where a statistical difference could be observed. Nevertheless, a previous study by Seitz et al. [14] confirmed that isokinetic knee extension exercises at 60°/s with a 6-s total contraction duration improved the maximal knee extension torque at 180°/s after the CA. The reason for this discrepancy may be the difference in the resistance training experience of the participants, which can affect the degree of PAPE [21]. The average age and resistance training experience of the SI group in the present study were 19.9 ± 1.6 and 3.5 ± 1.1 years, respectively (Table 1), while the average age of the participants in the Seitz et al. [14] study was 25.4 ± 3.9 years. Although their study did not state the years of resistance training experience, it can be presumed that the participants in their study had a longer resistance training history than those in the present study, which could explain the inconsistent results.

In contrast to SI, WI failed to display PAPE in any of the CAs (Fig 5B), and thus, we could not conclude whether the CAs performed with relatively short or long total contraction durations were optimal for WI. The fact that PAPE was observed in SI but not in WI after the 18-CA protocol may be due to the difference in the fatigue resistance against a high-intensity exercise. Miyamoto et al. [17] confirmed that after a 5-s maximal isometric knee extension exercise, the dynamic maximal knee extension torque increased 1 to 3 min later but not immediately after the CA. Interestingly, however, after 12 weeks of high-intensity resistance training, the potentiation effect of the voluntary performance was observed even immediately after the CA. The main reason for the earlier realization of PAPE after the intervention is that the individuals developed resistance to fatigue by chronic high-intensity resistance training, suppressing the remarkable fatigue associated with a high-intensity CA. In other words, this result means that the participants with higher muscle strength levels have a greater fatigue resistance to a high-intensity CA, and there is a high possibility that WI had a lower fatigue resistance to a high-intensity CA than that of SI in the present study. Therefore, it is likely that the significant fatigue associated with the CA was responsible for the lack of the potentiation of the voluntary performance in the 18-CA protocol for WI. In addition, contrary to our hypothesis, WI failed to enhance the voluntary performance after the CA with a relatively short total contraction duration (i.e., 6-CA). However, based on the individual data, we observed that for some participante (i.e., individuals numbered 15, 21, and 22), the CAs performed with short or moderate total contraction durations were suitable for maximizing PAPE (Fig 6). In comparison to a high-intensity CA with a relatively long contraction duration, a high-intensity CA with short or moderate total contraction duration may produce less fatigue and allow the potentiating effect to dominate even in WI with a relatively low fatigue resistance. Future studies should continue to investigate the optimal long total contraction duration required for WI to maximize PAPE. Furthermore, the enhancement in the maximal twitch torque was also absent in all protocols for WI. Hamada et al. [1] discovered that the magnitude of PAP depends on the muscle fiber type distribution, and participants with larger fast-twitch fibers evoked higher PAP. Considering that SI tend to have higher fast-twitch fiber content [19,20], they can trigger higher PAP through CA implementation. Fig 2 indicates that the higher the relative muscle strength, the larger the magnitude of PAP. Therefore, it is possible that the WI group, composed of participants with low muscle strength levels, did not significantly increase PAP due to the reduced number of fast-twitch fibers.

The present study has some limitations. First, the sample size of the SI and WI subgroups was small, and the participants with intermediate levels of relative muscle strength were not included in the subgroups to clearly distinguish the differences in muscle strength levels among the participants. A study with a larger sample size is necessary to confirm the present findings. Second, the athletic characteristics of the American football players who participated in this study may have influenced the results of this study. Specifically, since most of the game time in American football consists of intermittent high-intensity exercise, the participants in this study could have had higher fast-twitch fiber content, which is associated with the degree of PAP, than that of the athletes who require more endurance capacity (e.g., long-distance runners). Therefore, further studies should be conducted to determine whether similar results can be obtained in athletes with a high endurance capacity. Third, because the nature of isokinetic actions is different from that of isotonic actions, it may be difficult to directly apply the results to movements in sports and training. Further studies using isotonic exercise are required to apply the results of this study to the field. Fourth, the measures related to PAPE mechanisms that could help explain the mechanisms of the increase in voluntary performance observed after the 18-CA protocol, such as the central drive capacity, muscle temperature, and water content [26], were not examined in this study. Future studies should include these measures after a high-intensity CA with a long contraction duration. Lastly, the experimental conditions employed in this study did not allow us to determine whether the CAs performed with relatively short or long contraction durations are optimal for individuals with low muscle strength levels. Therefore, we hope that future studies with modification to the experimental conditions of this study (e.g., muscle contraction types, total contraction durations) will be conducted.

## Conclusion

The present study revealed that SI were able to enhance the voluntary performance only after the CA with a long contraction duration (i.e., 18-CA), whereas WI could not enhance the voluntary performance in all the CAs. The findings suggest that the CA with a relatively long contraction duration is optimal for maximizing PAPE in individuals with a high muscle strength. In contrast, we could not conclude whether the CAs performed with relatively short or long contraction durations are optimal for individuals with a low muscle strength. Based on the results of this study, it is suggested that coaches need to determine the strategies to maximize PAPE while considering the individual’s strength levels. Specifically, athletes with high relative muscle strength levels may likely prefer to select a CA performed with a relatively long contraction duration to induce the potentiation of the voluntary performance sufficiently. However, in this case, as PAP is not expected to be a primary mechanism for improving voluntary performance, CA focused on enhancing PAP may not always be effective to maximize PAPE. In contrast, athletes with a low relative muscle strength may need to complete chronic high-intensity resistance training as a first step to utilize PAPE. This may lead to the enhancement of fatigue resistance against high-intensity exercise, which is thought to contribute to the realization of PAPE. To improve the practical use of the current study, future studies should examine whether the results observed in this study are also observed when isotonic exercises are employed.

## Data Availability

All raw files are available from the excel database

## Acknowledgments

The authors would like to thank all participants for their cooperation and Editage (www.editage.jp) for the English language review.

## Notes

### Competing Interest Statement

The authors have declared that no competing interests exist.

### Funding Statement

The authors received no specific funding for this work.

### Author Declarations

This study was approved by the Ethics Review Committee of Osaka University of Health and Sport Sciences (approval number: 21-2). The study was conducted in conformity with the policy statement regarding the use of human participants by the Declaration of Helsinki.

## References

1. Hamada T, Sale DG, MacDougall JD, Tarnopolsky MA. Postactivation potentiation, fiber type, and twitch contraction time in human knee extensor muscles. J Appl Physiol (1985). 2000;88:2131–2137.doi: 10.1152/jappl.2000.88.6.2131.

2. Vandervoort AA, Quinlan J, Mccomas AJ. Twitch potentiation after voluntary contraction. ExpNeurol. 1983;81: 141–152. doi: 10.1016/0014-4886(83)90163-2.

3. Moore RL, Stull JT. Myosin light chain phosphorylation in fast and slow skeletal muscles in situ.Am J Physiol. 1984;247: C462–C471. doi: 10.1152/ajpcell.1984.247.5.C462.

4. Sweeney HL, Bowman BF, Stull JT. Myosin light chain phosphorylation in vertebrate striated muscle: regulation and function. Am J Physiol. 1993;264: C1085–C1095. doi:10.1152/ajpcell.1993.264.5.C1085.

5. Szczesna D. Regulatory light chains of striated muscle myosin. Structure, function and malfunction. Curr Drug Targets Cardiovasc Haematol Disord. 2003;3: 187–197. doi:10.2174/1568006033481474.

6. Tillin NA, Bishop D. Factors modulating post-activation potentiation and its effect on performance of subsequent explosive activities. Sports Med. 2009;39: 147–166. doi:10.2165/00007256-200939020-00004.

7. Baudry S, Duchateau J. Postactivation potentiation in a human muscle: effect on the load-velocity relation of tetanic and voluntary shortening contractions. J Appl Physiol (1985). 2007;103: 1318–1325. doi: 10.1152/japplphysiol.00403.2007

8. Bergmann J, Kramer A, Gruber M. Repetitive hops induce postactivation potentiation in triceps surae as well as an increase in the jump height of subsequent maximal drop jumps. PLOS ONE.2013;8: e77705. doi: 10.1371/journal.pone.0077705

9. Fukutani A, Takei S, Hirata K, Miyamoto N, Kanehisa H, Kawakami Y. Influence of the intensity of squat exercises on the subsequent jump performance. J Strength Cond Res. 2014;28: 2236–2243. doi: 10.1519/JSC.0000000000000409

10. Mitchell CJ, Sale DG. Enhancement of jump performance after a 5-RM squat is associated with postactivation potentiation. Eur J Appl Physiol. 2011;111: 1957–1963. doi: 10.1007/s00421-010-1823-x

11. Miyamoto N, Kanehisa H, Fukunaga T, Kawakami Y. Effect of postactivation potentiation on the maximal voluntary isokinetic concentric torque in humans. J Strength Cond Res. 2011;25:186–192. doi: 10.1519/JSC.0b013e3181b62c1d

12. Cuenca-Fernández F, Smith IC, Jordan MJ, MacIntosh BR, López-Contreras G, Arellano R, etal. Nonlocalized postactivation performance enhancement (PAPE) effects in trained athletes: apilot study. Appl Physiol Nutr Metab. 2017;42: 1122–1125. doi: 10.1139/apnm-2017-0217

13. Rassier DE, Macintosh BR. Coexistence of potentiation and fatigue in skeletal muscle. Braz JMed Biol Res. 2000;33: 499–508. doi: 10.1590/s0100-879×2000000500003

14. Seitz LB, Trajano GS, Dal Maso F, Haff GG, Blazevich AJ. Postactivation potentiation duringvoluntary contractions after continued knee extensor task-specific practice. Appl Physiol NutrMetab. 2015;40: 230–237. doi: 10.1139/apnm-2014-0377

15. Hamada T, Sale DG, MacDougall JD, Tarnopolsky MA. Interaction of fibre type, potentiationand fatigue in human knee extensor muscles. Acta Physiol Scand. 2003;178: 165–173. doi:10.1046/j.1365-201X.2003.01121.x

16. Jo E, Judelson DA, Brown LE, Coburn JW, Dabbs NC. Influence of recovery duration after apotentiating stimulus on muscular power in recreationally trained individuals. J Strength Cond Res. 2010;24: 343–347. doi: 10.1519/JSC.0b013e3181cc22a4.

17. Miyamoto N, Wakahara T, Ema R, Kawakami Y. Further potentiation of dynamic musclestrength after resistance training. Med Sci Sports Exerc. 2013;45: 1323–1330. doi:10.1249/MSS.0b013e3182874c0e

18. Seitz LB, de Villarreal ES, Haff GG. The temporal profile of postactivation potentiation is relatedto strength level. J Strength Cond Res. 2014;28: 706–715. doi: 10.1519/JSC.0b013e3182a73ea3.

19. Aagaard P, Andersen JL. Correlation between contractile strength and myosin heavy chainisoform composition in human skeletal muscle. Med Sci Sports Exerc. 1998;30: 1217–1222. doi:10.1097/00005768-199808000-00006.

20. Maughan RJ, Watson JS, Weir J. Relationships between muscle strength and muscle cross-sectional area in male sprinters and endurance runners. Eur J Appl Physiol Occup Physiol.1983;50: 309–318. doi: 10.1007/BF00423237.

21. Seitz LB, Haff GG. Factors modulating post-activation potentiation of jump, sprint, throw, andupper-body ballistic performances: A systematic review with meta-analysis. Sports Med.2016;46: 231–240. doi: 10.1007/s40279-015-0415-7.

22. Macintosh BR, Robillard ME, Tomaras EK. Should postactivation potentiation be the goal ofyour warm-up? Appl Physiol Nutr Metab. 2012;37: 546–550. doi: 10.1139/h2012-016.

23. Fukutani A, Miyamoto N, Kanehisa H, Yanai T, Kawakami Y. Potentiation of isokinetic torqueis velocity-dependent following an isometric conditioning contraction. Springerplus. 2013;2: 554.doi: 10.1186/2193-1801-2-554.

24. Prieske O, Maffiuletti NA, Granacher U. Postactivation potentiation of the plantar flexors doesnot directly translate to jump performance in female elite young soccer players. Front Physiol.2018;9: 276. doi: 10.3389/fphys.2018.00276.

25. Thomas K, Toward A, West DJ, Howatson G, Goodall S. Heavy-resistance exercise-inducedincreases in jump performance are not explained by changes in neuromuscular function. Scand JMed Sci Sports. 2017;27:35–44. doi: 10.1111/sms.12626

26. Blazevich AJ, Babault N. Post-activation potentiation versus post-activation performanceenhancement in humans: historical perspective, underlying mechanisms, and current issues. FrontPhysiol. 2019;10: 1359. doi: 10.3389/fphys.2019.01359.

27. De Ruiter CJ, De Haan A. Temperature effect on the force/velocity relationship of the fresh and fatigued human adductor pollicis muscle. Pflugers Arch. 2000;440: 163–170. doi:10.1007/s004240000284.

28. González-Alonso J, Quistorff B, Krustrup P, Bangsbo J, Saltin B. Heat production in humanskeletal muscle at the onset of intense dynamic exercise. J Physiol. 2000;524: 603–615. doi:10.1111/j.1469-7793.2000.00603.x.

29. Folland JP, Wakamatsu T, Fimland MS. The influence of maximal isometric activity on twitchand H-reflex potentiation, and quadriceps femoris performance. Eur J Appl Physiol. 2008;104:739–748. doi: 10.1007/s00421-008-0823-6.

30. Persechini A, Stull JT, Cooke R. The effect of myosin phosphorylation on the contractileproperties of skinned rabbit skeletal muscle fibers. J Biol Chem. 1985;260: 7951–7954. doi:10.1016/S0021-9258(17)39544-3.

31. Zimmermann HB, MacIntosh BR, Dal Pupo J. Does postactivation potentiation (PAP) increasevoluntary performance? Appl Physiol Nutr Metab. 2020;45: 349–356. doi: 10.1139/apnm-2019-0406.

